# Superficial White Matter Brain Alterations Discriminate Tinnitus in Older Adults

**DOI:** 10.1101/2025.07.28.25332324

**Authors:** Lázara Liset González Rodríguez, Simón Pedro San Martin Rubilar, Hernan Hernandez Larzabal, Carolina Delgado, Vicente Medel, Paul H. Délano, Pamela Guevara

## Abstract

Subjective tinnitus is the perception of sound in the absence of an external source, with a neurobiological basis that remains poorly understood. Importantly, tinnitus prevalence increases with aging, reaching up to 25 % in adults aged above 65 years. This study examines white matter tract alterations in older adults with tinnitus using diffusion magnetic resonance imaging. The research involved 96 individuals from the Chilean ANDES cohort, including 56 patients with tinnitus and 40 controls. Thirty-six deep white matter (DWM) and 84 superficial white matter (SWM) bundles were segmented. For each bundle, we extracted four diffusion tensor imaging metrics: axial diffusivity (AD), mean diffusivity (MD), radial diffusivity (RD), and fractional anisotropy (FA). With these features, we trained an Extreme Gradient Boosting classifier to predict tinnitus, achieving an AUC of 0.93, highlighting the relevance of AD. Key DWM bundles included the anterior arcuate fasciculus, inferior longitudinal fasciculus, and cingulum short fibers (right hemisphere), and the superior motor thalamic radiation (left hemisphere). Significant SWM bundles in the left hemisphere included the superior parietal, cuneus, lingual, superior temporal, caudal middle frontal, precentral, fusiform, postcentral, supramarginal, rostral middle frontal, and superior frontal regions. Tinnitus patients showed decreased AD, MD, and RD, and increased FA, suggesting microstructural reorganization. These changes may reflect adaptive or maladaptive plasticity. Increased FA could signal compensatory responses, while decreased AD might indicate axonal damage. Meanwhile, the decrease in MD and RD could indicate an increase in myelin integrity. This is the first study to investigate SWM bundle alterations in tinnitus patients.

## 1 Introduction

Subjective tinnitus is often described as the presence of a “phantom” sound in the absence of any identifiable sound source (Aha and Bankert 1996; Knipper et al. 2020). This type, as opposed to objective tinnitus, is more common and affects up to 16 million individuals annually in the United States (Shargorodsky et al. 2010). Its prevalence in the population can reach as high as 10-15 % (Bhatt et al. 2016), and it tends to become more prevalent with advancing age, often co-occurring with hearing impairment and cognitive disturbances. The highest incidence of tinnitus occurs among older adults, reaching up to 25 % in different cohorts in the United Kingdom (Stohler et al. 2019) and in the United States (Shargorodsky et al. 2010).

Tinnitus is among the most debilitating symptoms affecting mental health. The clinical repercussions of tinnitus are diverse and well-documented, including sleep disturbances, speech comprehension difficulties, frustration, and irritability, among others (Langguth et al. 2011). The presence of tinnitus is associated with a high risk of neuropsychiatric comorbidities such as anxiety and depression (Langguth et al. 2011; Bhatt et al. 2017). Also, cognitive disturbances can appear associated with tinnitus presence. Findings suggest that the distress caused by tinnitus may have a distinct impact on cognitive abilities, particularly affecting areas such as general or crystallized intelligence and executive functioning (Neff et al. 2021). Likewise, reaction time and processing speed can be detrimental in tinnitus subjects leading to cognitive inefficiency issues related to attentional difficulties (Malesci et al. 2021). Quality of life can be impacted by tinnitus specially in the elderly. Those subjects who present tinnitus during 5 years or more in later ages can show poorer scores in general physical, vitality and mental health domains compared to paired subjects without tinnitus (Oosterloo et al. 2021). Ultimately, neurocognitive detriment can be associated with tinnitus presence. Tinnitus severity has been identified as a potential independent predictor for mild cognitive impairment (MCI), indicating a possible intrinsic link between chronic tinnitus and cognitive decline. This association is of particular interest given the established connection between MCI and a higher risk of progressing to dementia (Lee et al. 2020).

Diffusion Magnetic Resonance Imaging (dMRI) has been widely used to detect changes in regions of white matter (WM) associated with tinnitus (Seydell-Greenwald et al. 2014; Benson et al. 2014; Gunbey et al. 2017; Alhazmi 2020; Jaroszynski et al. 2021; Khan et al. 2021; Eltabbakh and Nada 2023; Rosemann and Rauschecker 2023; Svobodová et al. 2024). This technique allows obtaining images that measure the diffusion of water molecules in brain tissues, providing insights into the microstructure of brain tissue. By utilizing diffusion information in each voxel of the image, it is possible to reconstruct a local diffusion model. Using this information alongside tractography algorithms (Basser et al. 2000; Smith et al. 2012; Malcolm et al. 2010; Tournier et al. 2012; Wasserthal et al. 2019), it is possible to compute the primary 3D pathways of axons within the cerebral white matter as a set of 3D curves, known as streamlines or brain fibers.

There are several approaches available for reconstructing local diffusion models. Among them, the Diffusion Tensor Imaging (DTI) model is the most commonly reported in tinnitus studies (Seydell-Greenwald et al. 2014; Benson et al. 2014; Gunbey et al. 2017; Alhazmi 2020; Jaroszynski et al. 2021; Khan et al. 2021). This model enables the calculation of diffusion indices such as fractional anisotropy (FA), mean diffusivity (MD), axial diffusivity (AD), and radial diffusivity (RD).

FA quantifies the degree of diffusion anisotropy and ranges from zero to one. Values close to zero indicate isotropic diffusion, as observed in cerebrospinal fluid, where diffusion occurs equally in all directions. In contrast, values close to one represent highly anisotropic diffusion, indicating high directional dependence, as found in coherent WM tracts. FA has been widely used to assess the integrity of cerebral white matter. MD quantifies the overall magnitude of water diffusion, with higher MD values denoting increased diffusivity. AD represents diffusion parallel to the principal axis of the tensor and is related to intrinsic axonal properties (Song et al. 2003). Lastly, RD measures diffusion perpendicular to the principal axis of the tensor and is associated with changes in myelination (Song et al. 2003). Among these metrics, FA and MD are the most commonly reported in tinnitus studies. Recently, the Neurite Orientation Dispersion and Density Imaging (NODDI) model (Zhang et al. 2012) has started to be used (Xu et al. 2025). This model provides three metrics: the intracellular volume fraction (ICVF), which estimates neurite density; the orientation dispersion index (OD), which quantifies the angular variation of neurite orientation; and the isotropic volume fraction (ISOVF), which represents the fraction of free water within the voxel. However, NODDI requires multi-shell dMRI data and is not yet widely adopted for clinical studies on tinnitus.

To study white matter alterations, brain fiber tracts are often classified into deep white matter (DWM) and superficial white matter (SWM) tracts. DWM fiber tracts are well described in the literature and are characterized by long-or medium-range, stable fibers with low inter-subject variability. In contrast, SWM fiber tracts have been less studied due to their higher inter-subject variability and their tendency to produce noisier tractography results (Guevara et al. 2020). However, recent advances in tractography have improved the ability to study these short association fibers (Schilling et al. 2025).

Most studies investigating tinnitus using dMRI have focused on microstructural differences between tinnitus patients and healthy controls. In both groups, participants may present comorbidities such as hearing loss. These studies typically include participants with mean ages below 60 years (Seydell-Greenwald et al. 2014; Benson et al. 2014; Gunbey et al. 2017; Alhazmi 2020; Jaroszynski et al. 2021; Khan et al. 2021; Eltabbakh and Nada 2023; Rosemann and Rauschecker 2023; Svobodová et al. 2024; Xu et al. 2025).

FA has been the most commonly reported metric for comparisons between patients and controls, followed by MD, and to a lesser extent, by other diffusion metrics. These measures have primarily been compared across anatomical regions using voxel-based morphometry (Ashburner and Friston 2000; Good et al. 2001), voxel-wise analysis (Smith et al. 2006a), fixel-based analysis (Raffelt et al. 2017), or tractography-based approaches (Jaroszynski et al. 2021).

The brain regions commonly investigated are those associated with auditory, attentional, and emotional functions (Seydell-Greenwald et al. 2014; Benson et al. 2014; Gunbey et al. 2017; Alhazmi 2020; Jaroszynski et al. 2021; Khan et al. 2021; Eltabbakh and Nada 2023; Rosemann and Rauschecker 2023; Svobodová et al. 2024). For example, the regions of interest (ROIs) include the central auditory pathway, particularly pathways from the lateral lemniscus, inferior colliculi, Heschl’s gyrus, planum temporale, medial geniculate body, and auditory cortex. Also examined are ROIs involving connections between the auditory system and the limbic system (amygdala, hippocampus, and parahippocampus), as well as association connections such as the arcuate fasciculus and the corpus callosum. Studies focusing on tractography-based fiber bundles are less common. Among these, the most recent study we identified investigated the acoustic radiation is from Jaroszynski et al. (2021).

The analysis methods for making comparisons are based on statistical approaches, such as statistical tests, regression, and correlation analysis. Previous studies using these analytical tools have reported variable results. For instance, in Seydell-Greenwald et al. (2014), an increase in FA was observed in the tinnitus group in the inferior colliculi (IC), and a positive correlation was found between FA and hearing loss. In contrast, Gunbey et al. (2017) found a decrease in FA in the IC, along with a negative correlation between FA and hearing loss. More recent studies also show a similar pattern: Jaroszynski et al. (2021) reported an increase in FA in the tinnitus group in the right inferior fronto-occipital fasciculus, while Khan et al. (2021) found a decrease in FA in the same fascicle. Recently, Xu et al. (2025) using Tract-Based Spatial Statistics (TBSS) (Smith et al. 2006b), reported reduced AD in the forceps minor and right corticospinal tract, as well as increased orientation dispersion (from NODDI) in the forceps minor in patients with tinnitus. They also found decreased structural connectivity between the right caudate and pericalcarine cortex, the right caudate and superior temporal gyrus, and between the left putamen and cuneus.

These different results may be explained by the fact that tinnitus presents a highly heterogeneous condition in terms of its audiological, behavioral, and psychological characteristics. Moreover, there is no consensus on which white matter dMRI metrics are the most effective for addressing tinnitus brain related changes. There is no comprehensive investigation of the characteristics of all DTI metrics in relation to tinnitus. Typically, studies have focused on characterizing deep white matter tracts, overlooking superficial white matter tracts. Additionally, most studies have been limited to comparing many DTI metrics in patients and controls and have not explored applying more advanced tools such as feature selection and combination methods. Some findings demonstrate that the combination of features can offer greater predictive power for the target variable. To our knowledge, this is the first study attempting to classify tinnitus and controls using superficial and deep white matter DTI metrics.

Our research goal is to explore the structural characteristics of both superficial and deep white matter in a group of older adults, categorizing them into individuals with tinnitus and those without it. This exploration utilizes the complete set of DTI measurements (axial diffusivity, AD; mean diffusivity, MD; radial diffusivity, RD; and fractional anisotropy, FA), alongside various classification algorithms, to examine the main characteristics of tinnitus subjects compared to their non-tinnitus peers.

## 2 Methods

In this work, we analyzed subjects with and without tinnitus from the Chilean ANDES cohort (Auditory and Dementia Study) (Belkhiria et al. 2019). Using dMRI data, we generated deterministic whole-brain tractography for each participant and identified the DWM and SWM fiber tracts for each individual using two fiber bundle atlases. Subsequently, we calculated the microstructural characteristics of each fiber bundle. Feature selection methods were applied, and classification models were trained using the microstructural metrics as predictor variables, and the presence of tinnitus as the outcome variable. Finally, we identified the fiber bundle microstructural features that were highly associated with the presence of tinnitus. Figure 1 summarizes the main procedures employed in this study, which are described in greater detail below.

**Fig. 1:**
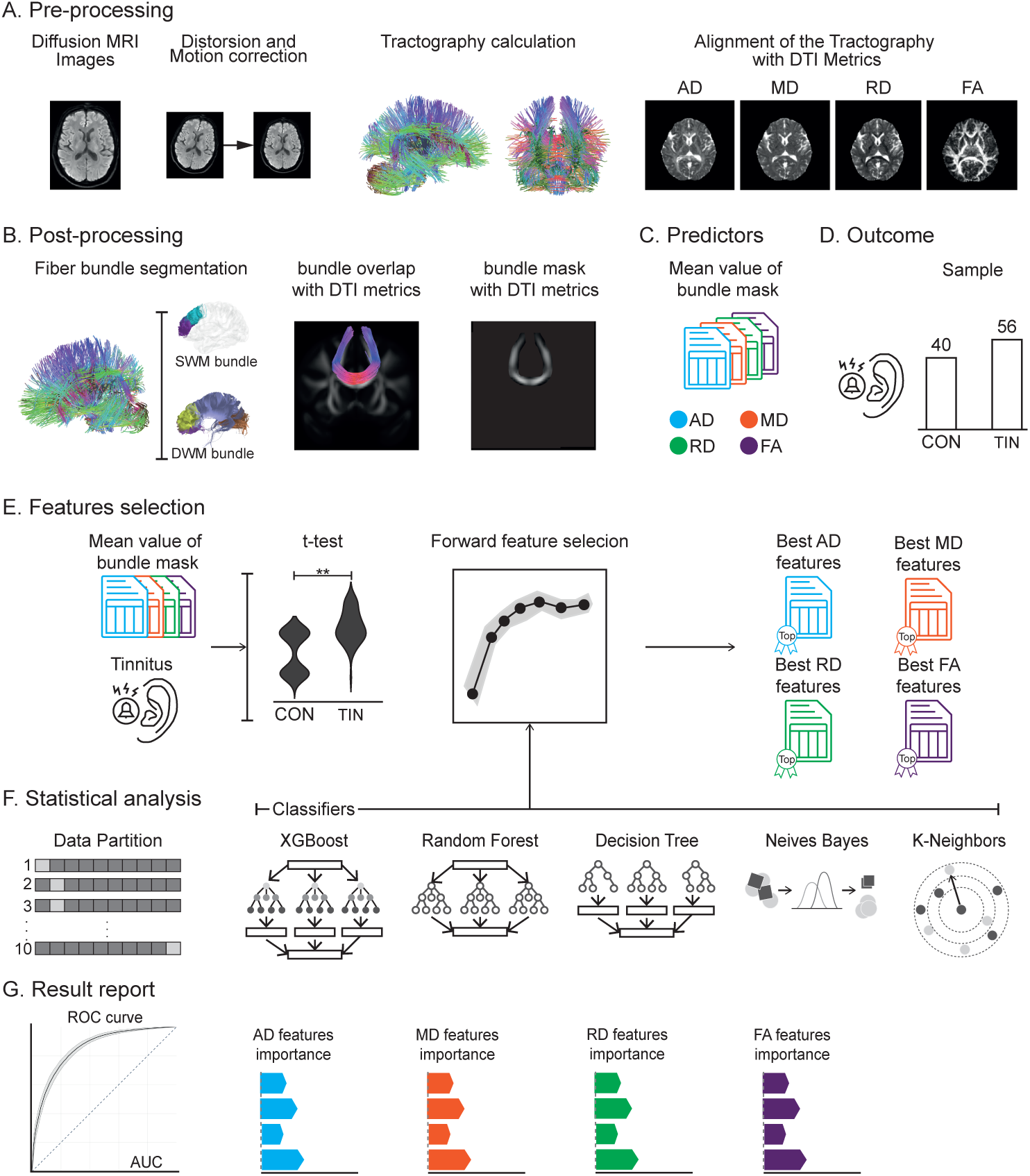
Processing pipeline for detecting the most influential fiber bundles in tinnitus prevalence. **A)** Preprocessing of diffusion MRI images involves motion and distortion correction. Deterministic tractography is calculated, and DTI (diffusion tensor images) are extracted and aligned to the subject’s space. **B)** Segmentation of fiber bundles using SWM and DWM bundle atlases. DTI masks are then extracted for all the bundles by intersecting segmented fiber bundles with DTI metrics (AD, MD, RD, and FA). **C)** Predictor extraction (mean DTI metrics for each bundle). We compute and store the average of the masks for each bundle. **D)** Outcome and sample size. The study aims to detect the presence of tinnitus (**TIN**) in a dataset of 96 participants (56 patients with tinnitus and 40 controls (**CON**). **E)** Multimethod feature selection approach. We employ a t-statistic test and forward feature selection to select the best features. The forward feature selection method used 5 classifiers (Decision Tree, XGBoost, Random Forest, Gaussian Naive Bayes, and K-Neighbors). **F)** Multimethod statistical analysis for participant classification. We applied 2a3multimethod approach using 5 classifiers. The classification was conducted through 10-fold cross-validation with 90% for training and 10% for testing. Additionally, hyperparameter optimization was performed for the best classifiers. **G)** Report on the classification results and the feature importance. We report the ROC curve, AUC, accuracy, precision, F1 score, sensitivity, and specificity.

### 2.1 Database

We used the ANDES (Auditory and Dementia Study) cohort dataset (Belkhiria et al. 2019). The study population was Chilean elderly individuals (aged 65 and older) who are non-demented and have a Mini-Mental State Examination (MMSE) score above 24. This study gathered clinical information on participants with varying degrees of age-associated auditory impairment, who had not used hearing aids before joining the study. Eligibility criteria included: maintained functional abilities as assessed by the Pfeffer Functional Activities Questionnaire, and magnetic resonance imaging (MRI) using a MAGNETOM Skyra 3-Tesla whole-body MRI Scanner (Siemens Healthineers, Erlangen, Germany) equipped with a head volume coil. The study excluded individuals with (i) hearing loss causes other than presbycusis, (ii) prior use of hearing aids, (iii) history of stroke or other neurological conditions, (iv) dementia and (v) significant psychiatric disorders. The University of Chile’s Clinical Hospital Ethics Committee approved all study procedures under the protocol OAIC 752/15. All methods were carried out in accordance with relevant guidelines and regulations. In line with the Declaration of Helsinki, informed consent was obtained from all subjects and/or their legal guardian(s). The database includes a total of 123 patients; however, our analysis focused on the 96 patients with available dMRI images: 56 with tinnitus and 40 controls (mean age: 73.92 years, standard deviation: ±5.6, 64% female). Table 1 provides a detailed overview of the clinical distribution of the participants included in our study.

**Table 1:** Clinical characteristics of analyzed patients. Clinical variables include diabetes, hypertension (HTA), dyslipidemia, and hearing loss. The “Total” column specifies the patient count for each variable. The “TIN” column indicates the number of patients with tinnitus, while the “CON” column denotes the number of patients without tinnitus.

| Clinical Variable | Total | TIN | CON |
| --- | --- | --- | --- |
| Diabetes | 26 | 15 | 11 |
| HTA | 63 | 40 | 23 |
| Dyslipidaemia | 47 | 27 | 20 |
| Hearing loss | 74 | 45 | 29 |

#### 2.1.1 Audiological data

Participants underwent an audiological evaluation by a skilled otolaryngology specialist. Hearing levels were assessed using air conduction from 0.125 to 8 kHz in both ears, and bone conduction thresholds were measured from 0.25 to 4 kHz to exclude conductive hearing loss. To further characterize hearing loss, the Pure Tone Average (PTA) was calculated for each participant, with the reported PTA representing the mean values across both ears. The distribution of hearing conditions among the subjects was as follows: 22.4% had normal hearing, 42.3% exhibited mild hearing loss, 31.4% had moderate hearing loss, and severe hearing loss was observed in only one subject (0.01%). This distribution indicates that mild hearing loss is the most prevalent condition among the study population, whereas moderate hearing loss and normal hearing are less common.

#### 2.1.2 dMRI acquisition

The diffusion imaging protocol utilized a specialized sequence designed for efficiency and participant comfort, featuring a 4 mm slice thickness and 30 diffusion directions. The sequence employed multi-slice scan mode and spin echo technique, with fast Echo Planar Imaging (EPI) in single-shot mode. Parameters included an echo time (TE) of 70/100 ms, a repetition time (TR) of 7200 ms, and a flip angle of 180*^◦^*. The acquisition matrix was 112 *×* 112, with a voxel size of 1.5 *×* 1.5 *×* 4.0 mm and a reconstruction voxel size of 2 mm. A 64-channel head coil ensured comprehensive brain coverage with a field of view (FOV) of 220. The total scan duration was 10 minutes and 17 seconds, making the setup rapid and effective for dMRI and tractography analysis.

### 2.2 dMRI data preprocessing

Diffusion MRI images were processed using DSI Studio software (Yeh et al. 2013). Firstly, eddy currents and motion corrections were applied. Subsequently, deterministic whole-brain tractography was computed using the DTI model and the following tracking parameters: angular threshold: 60*^◦^*, step size: 1 mm, smoothing: 0.5, minimum length: 30 mm, maximum length: 250 mm, and tract count: 1 million fibers. Then, the DTI model metrics were calculated, including axial diffusivity (AD), medial diffusivity (MD), radial diffusivity (RD), and fractional anisotropy (FA). Finally, non-linear deformation of the whole-brain tractography dataset into MNI space was performed. Figure 1A provides a visual overview of the entire preprocessing pipeline for diffusion MRI images.

#### 2.2.1 Fiber bundle segmentation and DTI metrics extraction

Our study focused on examining a large number of fiber bundles in both deep and superficial white matter, with particular emphasis on the latter, which has been less explored in tinnitus research. Fiber bundles were chosen using two comprehensive fiber bundle atlases: a DWM bundle atlas (Guevara et al. 2012) and a SWM bundle atlas (Román et al. 2017).

The selection of these atlases was based on their extensive coverage of white matter fascicles, allowing us to study both superficial and deep white matter comprehensively. The DWM bundle atlas contains 36 fascicles of deep white matter, and the SWM bundle atlas includes 93 fascicles of superficial white matter. This selection was made to ensure the study of a high number of fascicles in the brain. For further details on all analyzed fiber bundles, please refer to *Supplementary Tables S1* and *S2*.

To obtain the fiber bundles in each subject, we utilized the brain tractography dataset in MNI space, followed by the application of a fiber bundle segmentation algorithm based on a multi-subject atlas (Vázquez et al. 2019), available in the Phybers package (Gonźalez Rodríguez et al. 2024). Subsequently, masks of the bundles were constructed for each subject in the acquisition space. These masks were overlaid with images containing DTI metrics to extract the average value of each fiber bundle. This process is summarized in Figure 1B.

Consistently, all 36 fiber bundles in the DWM bundle atlas were detected in every participant of the study, whereas 84 out of the 93 bundles of the SWM bundle atlas were identified in all subjects. The difficulty in achieving a 100% detection rate for the SWM atlas in all subjects may be due to data quality or to the high variability among individual SWM fascicles. These fascicles exhibit greater structural and locational variability between individuals, than DWM bundles making some of them harder to detect consistently across all participants (Guevara et al. 2020; Schilling et al. 2025). This inherent variability and susceptibility to noise in the SWM fascicles contribute to the inability to identify all 93 bundles in every subject.

### 2.3 Feature extraction and classification from dMRI data

#### 2.3.1 Predictors

The predictors used in the analysis were the average DTI metrics extracted for each fiber bundle, as outlined in the previous step. These features were categorized into two groups according to the atlases employed. The first group consists of the measurements taken from the DWM bundles, while the second group encompasses DTI metrics from the SWM bundles. Additionally, we analyzed the behavior of four DTI metrics: AD, MD, RD, and FA. Based on these measures, four distinct feature matrices were constructed for each group (DWM and SWM bundles). In each matrix, columns represent the tract-based predictor variables, rows correspond to individual subjects, and the entries contain the respective DTI metric values. *Supplementary Table S1* provides a breakdown of the 36 DWM bundles extracted for the first group, while *Supplementary Table S2* offers details on the 83 assessed SWM bundles.

#### 2.3.2 Outcome

The dependent variable measured was the presence or absence of tinnitus. This variable was evaluated by the audiologist during the audiometer testing. Only those individuals reporting subjective tinnitus or controls without tinnitus were included in this study. Data from the ANDES cohort do not include tinnitus handicap inventory scores, thus we do not have a measure of tinnitus severity. Based on this assessment, participants were divided into two categories: those who reported any form of tinnitus over the course of the last year, and those who reported no presence of tinnitus.

#### 2.3.3 Feature selection

Feature selection was carried out using three different approaches: Statistical Feature Selection (SFS), Classification Feature Selection (CFS), and Merged Feature Selection (MFS). In the first two approaches, the four DTI metrics were analyzed separately for the DWM and SWM bundles. In contrast, the third approach considers metrics of the same type jointly across both DWM and SWM bundles. These approaches are described in detail below.

##### Statistical Feature Selection (SFS)

We applied t-test statistical analysis (Cohen 1988) and selected variables with a p-value below 0.05. This analysis was conducted separately for each group of fiber bundles and identified with the following names: SFS for DWM bundles (SFS-DWM) and SFS for SWM bundles (SFS-SWM).

##### Classification Feature Selection (CFS)

We employed five classification models in conjunction with forward sequential feature selection (Aha and Bankert 1996; Schooten et al. 2014). Specifically, we used the following classifiers: Decision Tree (Quinlan 1986; Rokach and Maimon 2005), Extreme Gradient Boosting (XGB) (Chen and Guestrin 2016), Random Forest (Breiman 2001; Liaw and Wiener 2001), Gaussian Naive Bayes (GNB) (Ontivero-Ortega et al. 2017; Jahromi and Taheri 2017), and K-Nearest Neighbors (KNN) (Cover and Hart 1967). These classifiers have proven useful in previous neuroscience studies (Douglas et al. 2013; Sarica et al. 2017; Wehenkel et al. 2018; Lebedev et al. 2014; Torlay et al. 2017; Yi et al. 2023; Ontivero-Ortega et al. 2017; Vrooman et al. 2007). We performed 10-fold cross-validation (Müller and Guido 2017), and feature selection was scored based on classification accuracy. As SFS, this analysis was performed separately for for DWM bundles (CFS-DWM) and SWM bundles (CFS-SWM).

##### Merged Feature Selection (MFS)

We combined the variables from the DWM and SWM bundles separately for each DTI metric, including only those that met the selection criteria of the first two approaches. This resulted in four feature sets representing the best-performing features for each DTI metric, as outlined below.

- ***MFS 1*** : SFS from the DWM bundles combined with SFS from the SWM bundles.
- ***MFS 2*** : CFS from the DWM bundles combined with CFS from the SWM bundles.
- ***MFS 3*** : SFS from the DWM bundles combined with CFS from the SWM bundles.
- ***MFS 4*** : CFS from the DWM bundles combined with SFS from the SWM bundles.

Consequently, the three feature selection methods resulted in a total of eight different feature sets.

#### 2.3.4 Classification models

For each feature set, we applied five classification models: Decision Tree, XGB, Random Forest, GNB, and KNN. The models were trained on 90% of the data and evaluated on the remaining 10% using 10-fold cross-validation. Performance was assessed based on the ROC curve and additional metrics, including precision, F1-score, sensitivity, specificity, and accuracy. Additionally, we report the feature importance weights for the best-performing models.

Hyperparameter tunning Before optimizing hyperparameters, we conducted searches to identify the best classification models for the three feature selection approaches outlined in the previous step. Model selection criteria were based on the area under the ROC curve (AUC). After identifying the top-performing models, we employed a Bayesian search (Feurer and Hutter 2019) with three-fold cross-validation and a 90/10% train-test split. For further details on the hyperparameters used, please refer to *Supplementary Tables S3*, *S4*, and *S5*.

## 3 Results

We evaluated the ability of DTI metrics to distinguish between tinnitus patients (n=56) and controls (n=40) using data from 84 SWM and 36 DWM fiber bundles. Three feature selection methods were applied: Statistical Feature Selection (SFS), Classification Feature Selection (CFS), and Merged Feature Selection (MFS). SFS and CFS each produced two feature sets per DTI metric (AD, MD, RD, FA), while MFS generated four, yielding a total of eight feature sets per metric. These sets were used as input for five classification models, resulting in 40 model evaluations per metric. In total, 160 classification models were executed across all four DTI metrics.

*Supplementary Table S6*. presents the top predictors identified by SFS for the DWM bundles, while *Supplementary Table S7* shows the corresponding results for the SWM bundles. *Supplementary Table S8* summarizes the best classifier, highest accuracy, and most influential predictors obtained through CFS for each DTI metric in the DWM bundles, and *Supplementary Table S9* provides the same information for the SWM bundles. The best classification performance was achieved using the MFS method, with results detailed in the following subsections.

### 3.1 Axial and mean diffusivities key indicators of tinnitus

The best classification performance was achieved for AD and MD using the MFS 3 feature selection method in both cases. In contrast, results for RD and FA were comparatively lower, with the highest performance obtained using the MFS 2 method.

The XGB model reached the highest AUC value of 0.93 for AD, as shown in Figure 2A (blue curve). Additional performance metrics are reported in Figure 2B (AD row), including an accuracy of 88.0%, precision of 95.5%, F1-score of 91.3%, sensitivity of 87.5%, and specificity of 89.3%. These values underscore the model’s strong performance, particularly in terms of sensitivity and specificity.

**Fig. 2:**
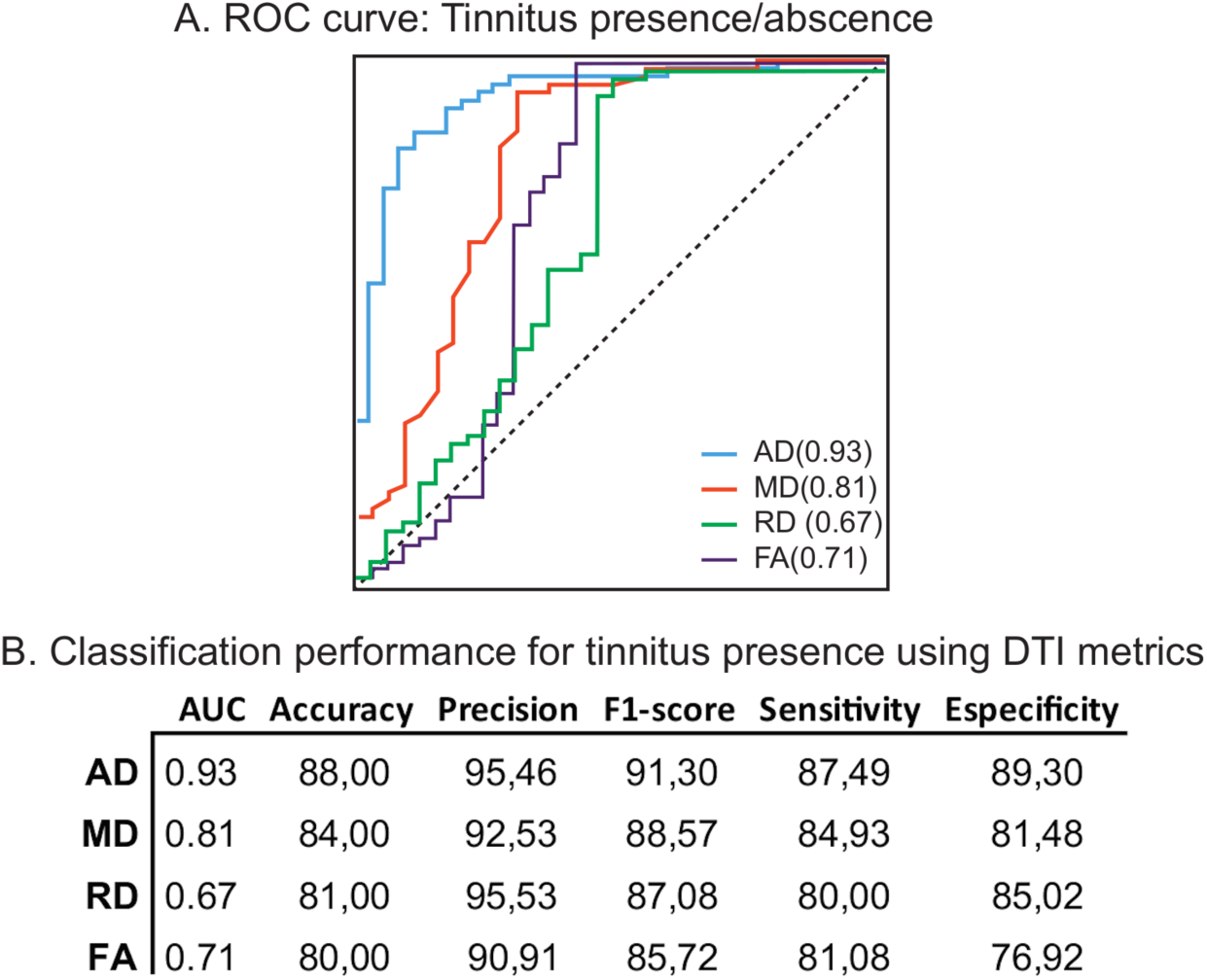
Best classification results for the four DTI metrics (AD, MD, RD, and FA) using the MFS method. **A)** ROC curves showing the overlap of the four curves: AD (blue), MD (orange), RD (green), and FA (purple). **B)** Classification performance for tinnitus presence using DTI metrics (rows) was evaluated for various metrics, including AUC, precision, F1 score, sensitivity, and specificity (columns).

The high sensitivity and specificity values observed in our analysis reflect the model’s strong ability to accurately identify both true positives and true negatives. High sensitivity indicates a low rate of false negatives, meaning the model reliably detects individuals with tinnitus, an essential aspect of this research, as it reduces the likelihood of undiagnosed cases. Likewise, high specificity minimizes false positives, ensuring that individuals without tinnitus are correctly classified, which enhances the reliability of our results. Overall, these metrics confirm the model’s effectiveness in discriminating between tinnitus patients and controls, thereby reinforcing the validity of our findings.

For MD, the best-performing model was Random Forest, which achieved an AUC of 0.81, as shown by the orange curve in Figure 2A. Figure 2B (MD row) reports the corresponding metrics: an accuracy of 84.0%, precision of 92.5%, F1-score of 88.6%, sensitivity of 84.9%, and specificity of 81.5%.

However, both RD and FA exhibit lower discriminative power between tinnitus patients and controls. For RD, the best performance is achieved with the GNB model, yielding a relatively low AUC of 0.67, as shown in green in Figure 2A. The corresponding metrics are: accuracy of 81.0%, precision of 95.5%, F1-score of 87.1%, sensitivity of 80.0%, and specificity of 85.0%, summarized in the RD row of Figure 2B.

For FA, the Random Forest model produces a similarly low AUC of 0.71, represented in purple in Figure 2A. The performance metrics include: accuracy of 75.0%, precision of 90.0%, F1-score of 87.0%, sensitivity of 86.0%, and specificity of 75.0%, as shown in the FA row of Figure 2B.

### 3.2 Relationship between merged SWM and DWM fiber bundle and tinnitus

We employed multiple feature selection methods, including Statistical Feature Selection, Classification Feature Selection, and Merged Feature Selection. Further details can be found in the Feature Selection Methods section (2.3.3). Among these, MFS yielded the best classification performance.

Combining features from both SWM and DWM fiber bundle atlases resulted in the highest classification accuracy across all four DTI metrics. The bar graph in Figure 3 displays the most influential fiber bundles contributing to these results. AD is shown in blue, MD in orange, RD in purple, and FA in green. Bundles are ranked in descending order of importance and grouped into DWM and SWM categories. SWM bundles are labeled according to the two ROIs they connect.

**Fig. 3:**
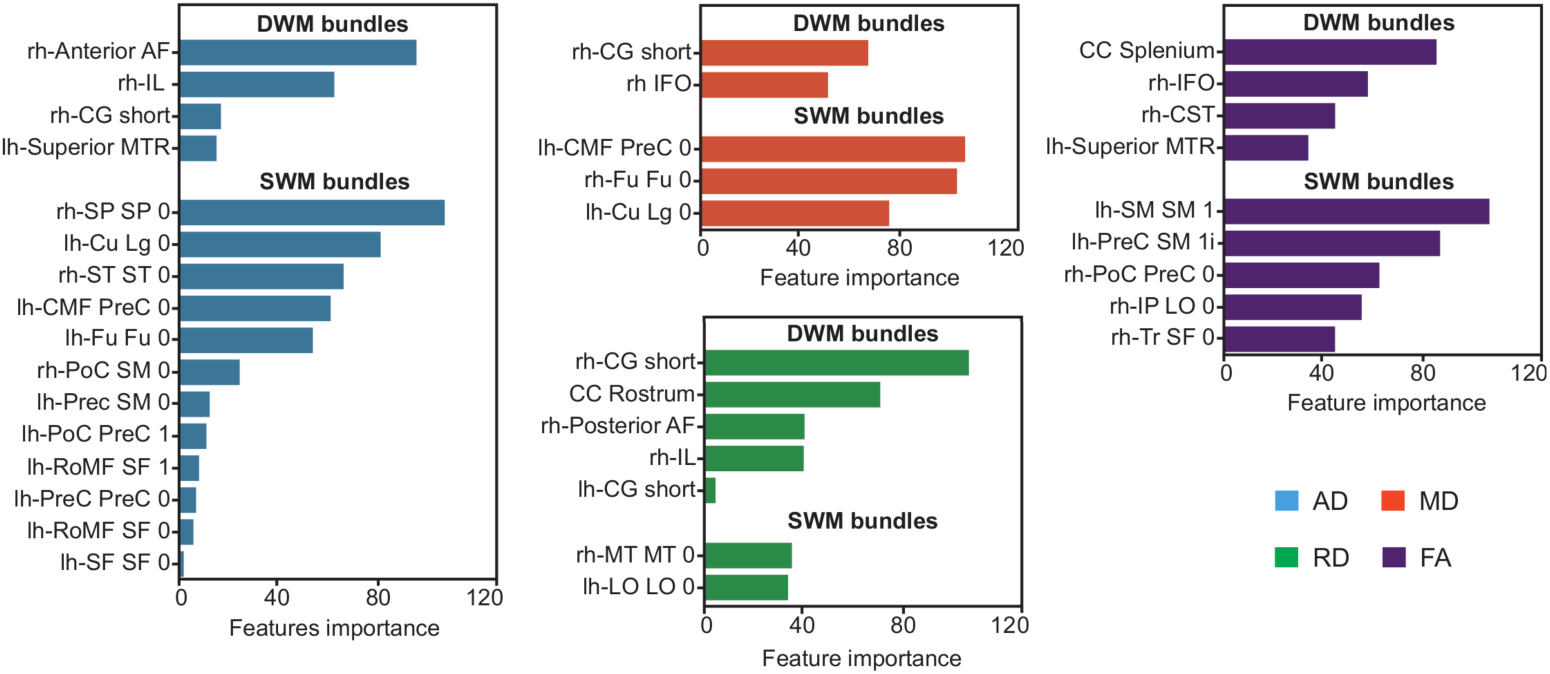
Most important fiber bundles for AD (blue), MD (orange), RD (purple), and FA (green) using the MFS method. The bundles are categorized into two groups, DWM and SWM bundles, and are also arranged in descending order. The prefixes **rh** and **lh** are utilized to denote the lateralization of the fiber bundles, **rh** (right hemisphere) and **lh** (left hemisphere). The abbreviations used for the fiber bundles are: **AF**, arcuate fasciculus; **IL**, inferior longitudinal fasciculus; **CG**, cingulum; **MTR**, motor thalamic radiations; **SP**, superior parietal; **Cu**, cuneus; **Lg**, lingual; **ST**, superior temporal; **CMF**, caudal middle frontal; **PreC**, precentral; **Fu**, fusiform; **PoC**, postcentral; **SM**, supramarginal; **RoMF**, rostral middle frontal; **SF**, superior frontal; **IFO**, inferior fronto-occipital fasciculus; **CC**, corpus callosum; **CST**, corticospinal tract; **IP**, inferior parietal; **LO**, lateral occipital; **Tr**, pars triangularis; and **MT**, middle temporal.

The nomenclature for fiber bundles follows a standardized pattern, where a bundle connecting **ROI1** and **ROI2** is labeled as **ROI1-ROI2-n**. The suffix “**n**” denotes the number of distinct connections between a given pair of ROIs in the atlas, where **n=0** represents the first connection, and **n=1** indicates a second one, if present. For example, a bundle connecting the postcentral (**PoC**) and supramarginal (**SM**) regions in the right hemisphere is labeled **rh-PoC-SM-0**. The prefix “**rh**” or “**lh**” denotes laterality, referring to the right or left hemisphere, respectively.

In our study, we identified connections involving the following ROIs: **MT** (middle temporal), **ST** (superior temporal), **PreC** (precentral), **SF** (superior frontal), **SM** (supramarginal), **CMF** (caudal middle frontal), **PoC** (postcentral), **RoMF** (rostral middle frontal), **Tr** (pars triangularis), **LO** (lateral occipital), **Fu** (fusiform), **IT** (inferior temporal), **Cu** (cuneus), **Lg** (lingual), **SP** (superior parietal), and **IP** (inferior parietal).

For AD (Figure 3, blue bars), the number of relevant DWM bundles is smaller compared to SWM bundles. The most important DWM bundles, ranked by their contribution, are: **rh-Anterior AF** (anterior arcuate fasciculus), **rh-IL** (inferior longitudinal fasciculus), **rh-CG short** (short cingulum fibers), and **lh-Superior MTR** (superior motor thalamic radiations).

In contrast, a larger set of short-range SWM bundles contributes to classification, including: **rh-SP-SP-0**, **lh-Cu-Lg-0**, **rh-ST-ST-0**, **lh-CMF-PreC-0**, **lh-Fu-Fu-0**, **rh-PoC-SM-0**, **lh-PreC-SM-0**, **lh-PoC-PreC-1**, **lh-RoMF-SF-1**, **lh-PreC-PreC-0**, and **lh-RoMF-SF-0**. Notably, the DWM bundles are predominantly located in the right hemisphere, whereas the SWM bundles are mostly found in the left hemisphere.

For MD (Figure 3, orange bars), fewer bundles contribute compared to AD. The most relevant DWM bundles are **rh-IFO** (inferior fronto-occipital fasciculus) and **rh-CG short**, while the key SWM bundles include **lh-CMF-PreC-0**, **rh-Fu-Fu-0**, and **lh-Cu-Lg-0**. Interestingly, the contributing DWM bundles are exclusively from the right hemisphere, while the SWM bundles show a predominance in the left hemisphere.

The main fiber bundles contributing to RD (Figure 3, green bars) include: **rh-CG short**, **CC Rostrum** (corpus callosum rostrum), **rh-Posterior AF** (posterior arcuate fasciculus), **rh-IL**, and **lh-CG short**. Most of these are located in the right hemisphere, with limited representation from the left. Notably, an interhemispheric connection, the **CC Rostrum**, also plays a significant role. The contribution of SWM bundles for RD is comparatively lower, with only two relevant bundles identified: **rh-MT-MT-0** and **lh-LO-LO-0**, located in the right and left hemispheres, respectively.

For FA (Figure 3, purple bars), the key DWM bundles include: **CC Splenium** (corpus callosum splenium), **rh-IFO**, **rh-CST** (corticospinal tract), and **lh-Superior MTR**. In this case, both hemispheres contribute, along with an interhemispheric connection via the **CC Splenium**. The SWM bundles associated with FA are distributed across both hemispheres and include: **lh-SM-SM-1**, **lh-PreC-SM-1**, **rh-PoC-PreC-0**, **rh-IP-LO-0**, and **rh-Tr-SF-0**.

### 3.3 Diffusivity decreases with the presence of Tinnitus

To further characterize differences in bundle-level DTI metrics between the tinnitus and control groups, box plots were generated and are presented in Figures 4 and 5.

**Fig. 4:**
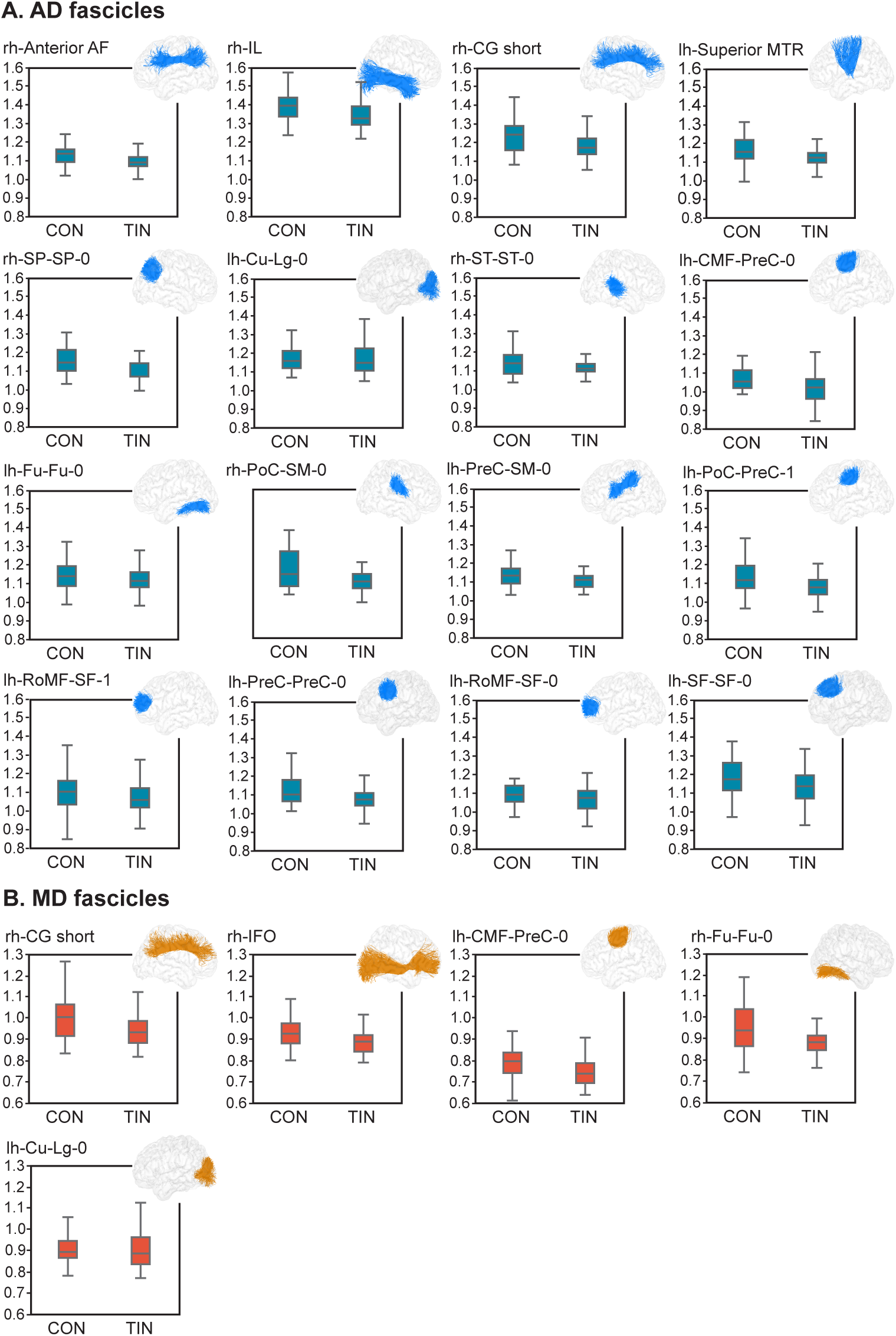
AD (axial diffusion) and MD (mean diffusion) results for the most important fiber bundles using the MFS method for tinnitus patients (**TIN**) and controls (**CON**). The prefixes **rh** and **lh** are utilized to denote the lateralization of the bundles, **rh** (right hemisphere) and **lh** (left hemisphere). **A)** denotes the DWM and SWM fiber bundles for AD. **B)** represents the DWM and SWM fiber bundles for MD. The abbreviations used for the fiber bundles are: **AF**, arcuate fasciculus; **IL**, inferior longitudinal fasciculus; **CG**, cingulum; **MTR**, motor thalamic radiations; **SP**, superior parietal; **Cu**, cuneus; **Lg**, lingual; **ST**, superior temporal; **CMF**, caudal middle frontal; **PreC**, pre-central; **Fu**, fusiform; **PoC**, postcentral; **SM**, supramarginal; **RoMF**, rostral middle frontal; **SF**, superior frontal; and **IFO**, inferior fronto-occipital fasciculus.

**Fig. 5:**
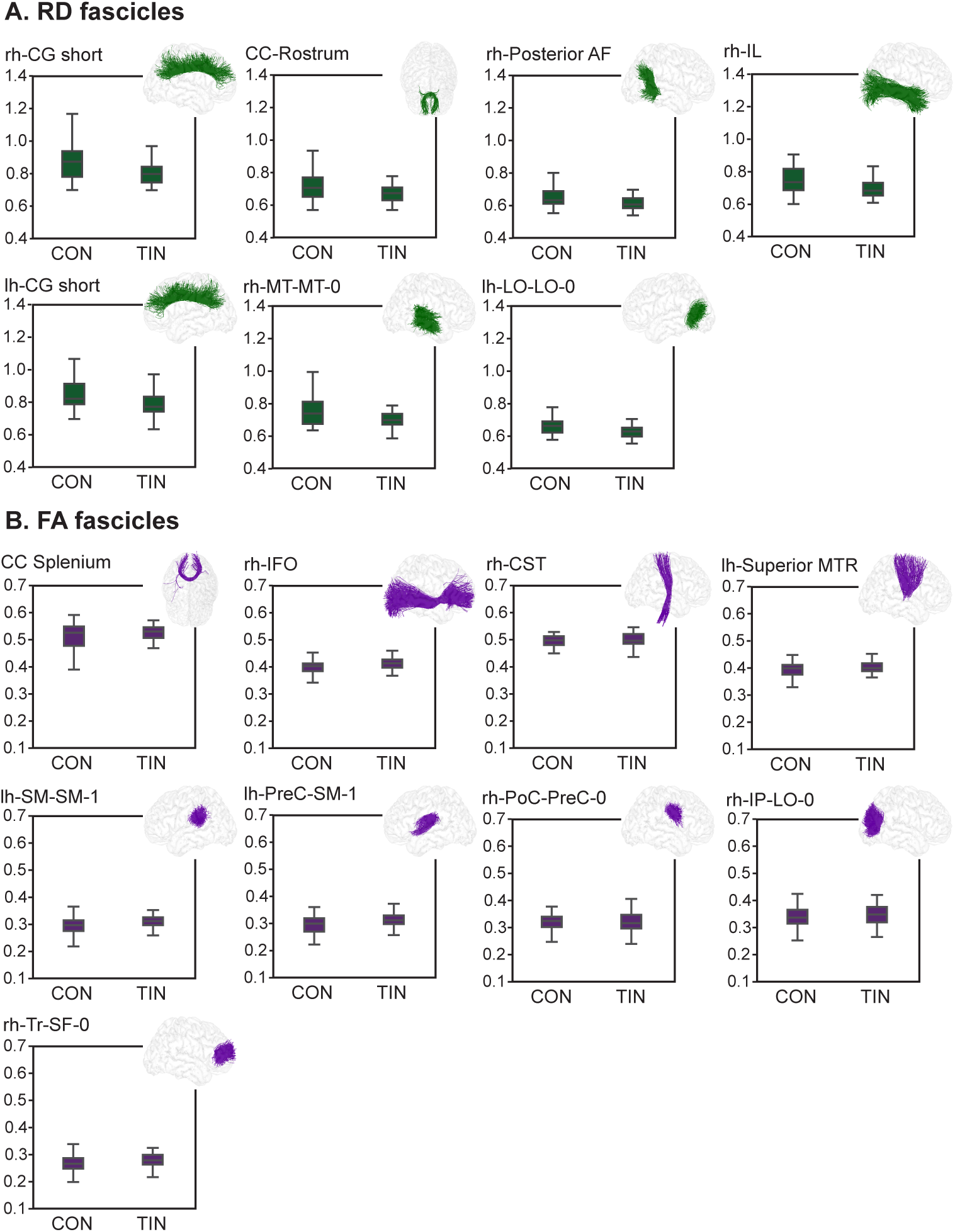
RD (radial diffusion) and FA (fractional anisotropy) in the most important fiber bundles using the MFS method for tinnitus patients (**TIN**) and controls (**CON**). The prefixes **rh** and **lh** are utilized to denote the lateralization of the fiber bundles, **rh** (right hemisphere) and **lh** (left hemisphere). **A)** denotes the long and short fiber bundles for RD. **B)** represents the long and short fiber bundles for FA. The abbreviations used for the fiber bundles are: **CG**, cingulum; **CC**, corpus callosum; **AF**, arcuate fasciculus; **IL**, inferior longitudinal fasciculus; **MT**, middle temporal; **LO**, lateral occipital; **IFO**, inferior fronto-occipital fasciculus; **CST**, corticospinal tract; **MTR**, motor thalamic radiations; **SM**, supramarginal; **PreC**, precentral; **PoC**, post-central; **IP**, inferior parietal; **Tr**, pars triangularis; and **SF**, superior frontal.

Figure 4A shows axial diffusivity, where a general decreasing trend is observed in the tinnitus group compared to controls. Similar patterns are seen for radial diffusivity and mean diffusivity, depicted in Figures 4B and 5A, respectively, with reduced values in the tinnitus group. In contrast, fractional anisotropy, shown in Figure 5B, exhibits an opposite trend: higher values in the tinnitus group and lower values in controls.

## 4 Discussion

Based on our objective to investigate structural alterations in superficial and deep white matter bundles in older adults with tinnitus, and leveraging a comprehensive set of DTI metrics along with multiple classification algorithms, this study provides key insights into the neuroanatomical characteristics of tinnitus. The combination of DWM and SWM features yielded the best classification performance, particularly for axial diffusivity and mean diffusivity. The XGB model demonstrated the highest effectiveness, achieving an AUC of 0.93 for AD. In addition, our analysis showed high accuracy, precision, and sensitivity, highlighting the robustness of the model in identifying individuals with tinnitus. These findings emphasize the potential of DTI-based machine learning approaches to detect subtle structural changes associated with tinnitus and support the development of more effective diagnostic and treatment strategies for this population.

The findings indicate that axial diffusivity and mean diffusivity possess strong discriminatory power for distinguishing between tinnitus patients and controls. The merged feature selection method proved to be the most effective, leading to the highest classification performance for both AD and MD. These results are supported by high AUC values, highlighting the robustness of the classification models. In contrast, radial diffusivity and fractional anisotropy exhibited comparatively lower discriminative capacity. Despite the use of multiple classification algorithms, AUC values for RD and FA remained modest, suggesting these metrics may be less reliable for differentiating tinnitus in this cohort.

Analysis of the most influential fiber bundles further underscores the value of combining SWM and DWM information to better understand the structural correlates of tinnitus. This approach revealed distinct anatomical patterns among the key predictors: DWM bundles were predominantly located in the right hemisphere, suggesting asymmetric alterations in white matter integrity, while SWM bundles showed a stronger presence in the left hemisphere. These findings offer additional insight into the spatial distribution of microstructural changes associated with tinnitus.

Our most significant finding was a widespread decrease in axial diffusivity in tinnitus patients across multiple white matter bundles. Among the most relevant DWM bundles identified in the right hemisphere were the **rh-Anterior AF** (anterior arcuate fasciculus), **rh-IL** (inferior longitudinal fasciculus), and **rh-CG short** (short cingulate fibers); in the left hemisphere, the key bundle was the **lh-Superior MTR** (superior motor thalamic radiations). The functional roles of these structures provide insight into their possible involvement in tinnitus. In the right hemisphere, the arcuate fasciculus is implicated in visuospatial processing and aspects of language such as prosody and semantics. The inferior longitudinal fasciculus connects the ipsilateral occipital and temporal lobes, forming long association fibers that link visual areas with the amygdala and hippocampus (Catani and Thiebautdeschotten 2008). It plays a role in facial recognition, visual perception, reading, visual memory, and language processing. The cingulum, a medium-sized fiber tract that runs along the cingulate sulcus around the corpus callosum, connects different parts of the cingulate cortex and various brain lobes (Catani and Thiebautdeschotten 2008). As part of the limbic system, it is involved in attention, memory, and emotional regulation. Finally, the internal capsule and corona radiata contain ascending fibers from the thalamus to the cerebral cortex and descending fibers from the frontoparietal cortex to subcortical nuclei (e.g., basal ganglia and brainstem nuclei) and the spinal cord (Catani and Thiebautdeschotten 2008). This complex projection system forms the neuroanatomical backbone of perceptual, motor, and higher-order cognitive functions.

Among the most relevant SWM bundles, those located in the right hemisphere include: **rh-SP-SP-0** (superior parietal–superior parietal), **rh-ST-ST-0** (superior temporal–superior temporal), and **rh-PoC-SM-0** (postcentral–supramarginal). In the left hemisphere, the key bundles are: **lh-Cu-Lg-0** (cuneus–lingual), **lh-CMF-PreC-0** (caudal middle frontal–precentral), **lh-Fu-Fu-0** (fusiform–fusiform), **lh-PreC-SM-0** (precentral–supramarginal), **lh-PoC-PreC-1** (postcentral–precentral), **lh-RoMF-SF-1** and **lh-RoMF-SF-0** (rostral middle frontal–superior frontal), and **lh-PreC-PreC-0** (precentral–precentral).

SWM bundles connect adjacent cortical regions and play a crucial role in local information integration. In the context of tinnitus, the involvement of specific cortical areas connected by these bundles may shed light on the neural mechanisms underlying the perception and experience of tinnitus. For example, among the mentioned brain regions, such as the superior parietal, cingulum-lingual, and superior temporal, there are areas involved in attention, auditory perception, and memory. The superior parietal is associated with spatial perception and selective attention, while the cingulum-lingual and superior temporal are involved in auditory processing and multisensory integration.

The analysis also reveals alterations in diffusivity associated with tinnitus. The box plots (Figures 4 and 5) show a consistent pattern of reduced diffusivity, particularly in AD, MD, and RD, among tinnitus patients compared to controls. In contrast, FA displays an opposing trend, with increased values observed in the tinnitus group. Previous studies have reported that decreased FA is typically associated with hearing loss in control subjects, whereas tinnitus patients with hearing loss tend to exhibit elevated FA values (Khan et al. 2021). These findings suggest that the observed changes in white matter integrity may reflect a compensatory neuroplastic response to the auditory deficit.

To our knowledge, this is the first study to examine SWM bundles in patients with tinnitus. However, prior work has investigated DWM bundles, particularly those related to the acoustic radiations (Jaroszynski et al. 2021). Our findings align with those of (Jaroszynski et al. 2021), who also reported increased FA in the inferior fronto-occipital fasciculus (IFO) of the right hemisphere in tinnitus patients. The IFO is a ventral associative fiber tract that connects the ventral occipital lobe with the orbitofrontal cortex. It plays a critical role in language processing, attentional control, affective regulation, and visual recognition systems (Catani and Thiebautdeschotten 2008). Moreover, it has been suggested that direct connections between the IFO and Heschl’s gyrus may contribute to the perception of tinnitus (Ferńandez et al. 2020). This finding may help explain the cognitive and specifically, attentional difficulties in tinnitus subjects.

Despite the significant findings and contributions of this study, several limitations should be acknowledged. First, we opted to compute the average of DTI metrics along the entire length of each fascicle. In contrast, more recent approaches, such as that of Jaroszynski et al. (2021), segment each tract into 100 parts and calculate average metrics for each segment individually. This method allows for more precise localization of microstructural alterations along the tract and facilitates the exclusion of the tract ends, which are often affected by noise. Additionally, our analysis relied exclusively on DTI measures, without incorporating more advanced models such as fixel-based analysis, which has been applied in other recent studies (Rosemann and Rauschecker 2023; Svobodová et al. 2024). These methods provide improved characterization of white matter microstructure, particularly in regions with crossing fibers. Addressing this limitation may require the adoption of more sophisticated diffusion models, such as constrained spherical deconvolution (CSD), which could entail the reacquisition of diffusion MRI data.

Regarding the study cohort, it was composed of older adults, which may limit the generalizability of the findings to other age groups or populations. Furthermore, we acknowledge the need to increase the sample size and to consider longitudinal follow-up of participants to better understand the progression of tinnitus-related white matter changes.

## 5 Conclusion

The results of this study provide valuable insights into the discriminatory potential of DTI metrics (AD, MD, RD, and FA) for distinguishing between patients with tinnitus and controls, all of whom were older adults with potential comorbidities. A large number of fiber bundles were examined, including 36 from deep white matter and 84 from superficial white matter, the latter of which has been less extensively studied (Guevara et al. 2020; Schilling et al. 2025). This comprehensive analysis offers a detailed perspective on the structural connectivity underlying tinnitus. The analysis was conducted through the application of various feature selection methods and classification algorithms. XGBoost emerged as the top-performing classifier, particularly when using a feature selection strategy that combined fiber tracts from both the DWM and SWM atlases. This underscores the important contribution of SWM bundles to the characterization of tinnitus-related white matter alterations.

Our study sheds new light on the alterations of SWM and DWM bundles in patients with tinnitus. This research underscores the importance of these alterations and highlights the complexity of tinnitus as a condition influenced by widespread neuroplastic changes.

While previous research has primarily focused on central auditory pathways and cortical regions, the potential role of SWM bundles has been largely understudied. By presenting the first comprehensive analysis of SWM bundle alterations in tinnitus patients, our study helps bridge this gap. This novel perspective is particularly important, as SWM bundles are essential for intracortical communication. Disruptions in these short-range connections may contribute to aberrant neural activity and, ultimately, to the perception of phantom sounds.

The findings suggest several mechanisms through which SWM bundles may contribute to the development of tinnitus. The observed decrease in AD, MD, and RD, coupled with an increase in FA, points to microstructural reorganization within these fiber pathways. Such alterations may reflect either adaptive or maladaptive plasticity in response to auditory deprivation or damage. In particular, the increased FA may indicate a compensatory mechanism, whereby remaining intact fibers become more directionally organized to preserve auditory processing efficiency despite peripheral deficits. In contrast, the reductions in AD, MD, and RD could suggest demyelination or axonal degeneration, potentially disrupting normal auditory signal transmission and contributing to the persistence of tinnitus.

Future research can leverage advanced neuroimaging techniques to further elucidate the relationship between SWM bundles and tinnitus. High-resolution diffusion MRI, in combination with probabilistic tractography algorithms, can enable more precise mapping of both SWM and DWM bundles. Additionally, techniques such as fixel-based analysis (Raffelt et al. 2017), neurite orientation dispersion and density imaging (NODDI) (Zhang et al. 2012), and other multi-compartment models can offer deeper insight into microstructural properties by disentangling contributions from axonal density, fiber orientation, and myelin content. Complementary functional modalities, such as functional MRI (fMRI) and magnetoencephalography (MEG), can help relate these structural abnormalities to functional connectivity patterns and altered neural oscillations associated with tinnitus.

Understanding the structural alterations in SWM and DWM bundles opens new avenues for the development of targeted treatment strategies for tinnitus. Therapeutic interventions could aim to restore or enhance the integrity of affected white matter pathways. For instance, non-invasive brain stimulation techniques, such as transcranial magnetic stimulation (TMS) or transcranial direct current stimulation (tDCS), could be tailored to enhance neuroplasticity and repair damaged fiber bundles. Additionally, pharmacological approaches targeting neuroinflammation and promoting remyelination may prove beneficial. Rehabilitation programs incorporating auditory training and cognitive behavioral therapy could also be adapted to leverage the plasticity of these pathways, potentially reducing the perception of tinnitus.

In conclusion, this study highlights the critical role of both superficial and deep white matter tracts in the pathophysiology of tinnitus, addressing a significant gap in the existing literature. The structural alterations identified through diffusion MRI provide a foundation for advancing our understanding of the complex neural mechanisms underlying tinnitus. These findings also pave the way for the development of innovative, targeted treatment strategies aimed at alleviating the burden of this challenging condition.

## Data Availability

All data produced in the present study are available upon reasonable request to the authors.

## Supplementary information

A supplementary file is provided containing detailed information on the names of the analyzed fiber bundles, the results of the feature selection procedures and classification models,.

## Acknowledgements

ANDES Database. Presbycusis Neuroimaging Data were contributed by the collaborative research team at the Neuroscience Department, Facultad de Medicina, Universidad de Chile, Santiago, Chile; and the Otolaryngology Department, Hospital Cĺınico de la Universidad de Chile, Santiago, Chile. The study (Principal Investigators: Paul H. Delano and Carolina Delgado) was supported by the Proyecto Anillo ACT1403, ANID BASAL AFB240002, FONDEF ID20I10371, FONDECYT grants (1221696, 1220607, 3230557) and Fundacíon Guillermo Puelma. Data collection encompassed comprehensive audiometric evaluations, neuropsychiatric assessments, and high-resolution magnetic resonance imaging (MRI) at 3 Tesla, facilitating the exploration of structural brain alterations associated with presbycusis and its impact on functional impairment in older adults.

## Ethics approval and consent to participate

This study was performed in line with the principles of the Declaration of Helsinki. Approval was granted by the Ethics Committee of Clinical Hospital of the University of Chile (Date: 12 November 2014/No OAIC752/15). Informed consent was obtained from all individual participants included in the study.

## Author contribution

L.L.G.R. analyzed the MRI data, conceived and conducted the experiment(s), analysed the results, drafted the manuscript; S.P.M.R. collected dMRI and audiological data, analysed the results, drafted the manuscript; H.H.L. conceived the experiment(s), drafted the manuscript; C.D. collected dMRI and audiological data, obtained funding the study; V.M. collected dMRI and audiological data; P.H.D. collected dMRI and audiological data, obtained funding and designed the study; P.G. obtained funding and designed the study. All authors reviewed the manuscript.

## Data and code availability

The data and code that support the findings of this study are available from the corresponding authors upon reasonable request.

## Supplementary materials

**Supplementary Table S1.** Names of the fiber bundles in the deep white matter (DWM) bundle atlas. The bundles without an asterisk correspond to tracts present in both hemispheres (right and left), while bundles marked with an asterisk represent interhemispheric connections.

| Names of the DWM bundles |  |
| --- | --- |
| Direct Arcuate fasciculus (Direct AR) | Cingulate Long fibers (CG Long) |
| Posterior Arcuate fasciculus (Posterior AR) | Cingulate Short fibers (CG Short) |
| Anterior Arcuate fasciculus (Anterior AR) | Cingulate Temporal fibers (CG Temporal) |
| Anterior Thalamic Radiations (Anterior TR) | Uncinate fasciculus (UF) |
| Superior Motor Thalamic Radiations (Superior Motor TR) | Corticospinal tract (CST) |
| Superior Parietal Thalamic Radiations (Superior Parietal TR) | Fornix fasciculus (FF) |
| Posterior Thalamic Radiations (Posterior TR) | Corpus callosum Body* (CC Body) |
| Inferior Thalamic Radiations (Inferior TR) | Corpus callosum Genu* (CC Genu) |
| Inferior Longitudinal fasciculus (IL) | Corpus callosum Rostrum* (CC Rostrum) |
| Inferior Fronto-Occipital fasciculus (IFO) | Corpus callosum Splenium* (CC Splenium) |

**Supplementary Table S2.** Names of the fiber bundles in the superficial white matter (SWM) bundle atlas. SAM bundles names are derived from the two regions of interest (ROIs) they connect, according to the anatomical Desikan-Killiany atlas. The regions in the Desikan-Killiany atlas include: Middle Temporal (MT), Superior Temporal (ST), Precentral (PreC), Superior Frontal (SF), Supramarginal (SM), Insula (Ins), Caudal Middle Frontal (CMF), Pars Opercularis (Op), Postcentral (PoC), Rostral Middle Frontal (RoMF), Pars Triangularis (Tr), Lateral Occipital (LO), Fusiform (Fu), Inferior Temporal (IT), Cuneus (Cu), Lingual (Lg), Precuneus (PreCu), Superior Parietal (SP), Inferior Parietal (IP), and Lateral Orbitofrontal (LOrF). The nomenclature for the bundles follows a standard pattern, where a bundle connecting ROI1 and ROI2 is named as ROI1-ROI2-n. The ‘n’ number is assigned within the range of 0 to 3, indicating the specific connection among for the pair of ROIs. Names without an asterisk correspond to bundles present in both hemispheres (right and left), while those marked with a single asterisk represent connections only in the left hemisphere, and those with a double asterisk represent connections only in the right hemisphere. Names marked in red correspond to bundles that were not detected in some subjects; therefore, they were excluded from the analysis.

| Names of the SWM bundles |  |  |  |
| --- | --- | --- | --- |
| MT-MT-0 | CMF-Op-0 | IT-IT-1* | SM-SM-1 |
| MT-MT-1 | Tr-RoMF-0 | PreCu-PreCu-0* | SM-SM-2 |
| MT-MT-1* | Tr-SF-0 | PreC-SF-0 | IP-SP-0 |
| MT-MT-0** | Tr-Tr-0** | PreC-SM-1 | IP-LO-0** |
| MT-ST-0 | Tr-Ins-0** | PreC-SM-0 | IP-IP-0** |
| RoMF-RoMF-0 | Tr_SF-1** | PreC-PreC-0** | Fu-Fu-0 |
| RoMF-RoMF-1 | LO_LO-0 | PreC-Ins-0** | Fu-IT-0 |
| RoMF-SF-1 | LO-LO-1 | PoC-PreC-0 | Fu-Fu-1* |
| RoMF-SF-0 | LO-LO-2* | PoC-PreC-1 | SF-SF-0* |
| RoMF-RoMF_0** | ST-ST-0 | PoC-PreC-2 | SF-SF-2** |
| RoMF-RoMF-1** | ST-ST-1* | PoC-PreC-3 | SF-SF-1** |
| RoMF-SF-0** | LOrF-LOrF-0 | PoC-SM-0 | Op-SF-0 |
| CMF-CMF-0 | LOrF-LOrF-1** | PoC-PoC-1** | SP-SP-0 |
| CMF-PreC-0 | PreCu-PreCu-0** | PoC-PreC-1** | SP-SP-0** |
| CMF-PreC-1 | IT-IT-0* | SM-SM-0 | Cu-Lg-0* |

**Supplementary Table S3.** Hyperparameters used for the XGBoost model. The first column lists the hyperparameter names, and the second column contains the range of values explored for each hyperparameter.

| **XGBoost hyperparameters** | **Values** |
| --- | --- |
| Number of Estimators | 10, 50, 100, 200 |
| Learning Rate | None, 10, 20, 30 |
| Maximum Depth of Trees | 2, 5, 10 |
| Subsample Ratio of Training Data | 1, 2, 4 |
| Subsample Ratio of Columns for Building Trees | "auto", "sqrt2", "log2" |
| L1 Regularization Term on Weights | True |
| L2 Regularization Term on Weights | "gini", "entropy" |
| Minimum Loss Reduction Required to Make a Further Partition on a Leaf Node | None, "balanced" |

**Supplementary Table S4.**
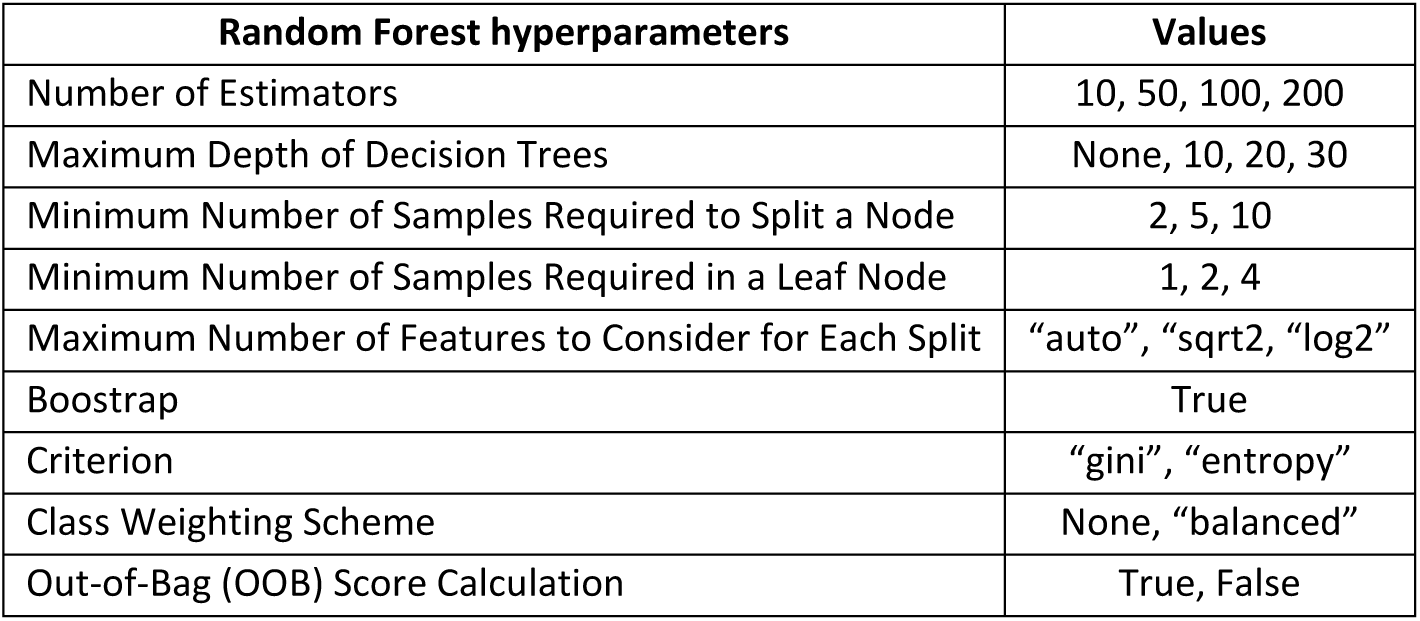
Hyperparameters used for the Random Forest model. The first column lists the hyperparameter names, and the second column contains the range of values explored for each hyperparameter.

**Supplementary Table S5.** Hyperparameters used for the Gaussian Naive Bayes model. The first column lists each hyperparameter name, and the second column contains the range of values explored for each.

| **GaussianNB Hyperparameters** | **Values** |
| --- | --- |
| Variance Smoothing (var_smoothing) | 1e-9, 1e-8, 1e-7, ..., 1e-5 (log-uniform range) |

**Supplementary Table S6.** T-test statistics for the fiber bundles in DWM for diffusion tensor model metrics: AD, axial diffusivity; MD, mean diffusivity; RD, radial diffusivity; and FA, fractional anisotropy. Only bundles that demonstrate statistical significance (p-value < 0.05, uncorrected) and an effect size above 0.5 (Cohen’s d) are included. The prefixes rh and lh indicate the laterality, rh (right hemisphere) and lh (left hemisphere).

| AD |  |  |
| --- | --- | --- |
| Bundle | p-value | d-Cohen |
| rh-Anterior AF | 0.002782 | 0.64 |
| lh-CST | 0.004487 | 0.60 |
| rh-CG Short | 0.007489 | 0.57 |
| lh-CG Short | 0.008291 | 0.56 |
| rh-IL | 0.011530 | 0.53 |
| lh-Anterior AF | 0.012007 | 0.53 |
| lh-Superior MTR | 0.014099 | 0.52 |

| MD |  |  |
| --- | --- | --- |
| Bundle | p-value | d-Cohen |
| lh-CST | 0.001931 | 0.66 |
| lh-Posterior AF | 0.002659 | 0.64 |
| rh-CG Short | 0.004995 | 0.60 |
| lh-CST | 0.008286 | 0.56 |
| rh-IL | 0.008673 | 0.55 |
| lh-CG Short | 0.012528 | 0.53 |
| lh-Superior MTR | 0.013402 | 0.52 |
| rh-Anterior AF | 0.013605 | 0.52 |
| rh-Posterior AF | 0.015222 | 0.51 |

| RD |  |  |
| --- | --- | --- |
| Bundle | p-value | d-Cohen |
| lh-Posterior AF | 0.001702 | 0.67 |
| rh-CG Short | 0.004746 | 0.60 |
| lh-CST | 0.006277 | 0.58 |
| rh-Posterior AF | 0.008777 | 0.55 |
| rh-IL | 0.009455 | 0.55 |
| CC Body | 0.010651 | 0.54 |
| CC Rostrum | 0.010716 | 0.54 |
| lh-CG Long | 0.013951 | 0.52 |
| CC Genu | 0.014947 | 0.51 |
| lh-Superior MTR | 0.018094 | 0.50 |

| FA |  |  |
| --- | --- | --- |
| Bundle | p-value | d-Cohen |
| CC Rostrum | 0.001767 | 0.67 |
| lh-Posterior AF | 0.002653 | 0.64 |
| lh-UN | 0.004475 | 0.60 |
| rh-Posterior AF | 0.006278 | 0.58 |
| lh-IL | 0.007186 | 0.57 |
| CC Genu | 0.008343 | 0.56 |
| CC Body | 0.013951 | 0.52 |
| lh-CG Long | 0.016045 | 0.51 |
| rh-CG Short | 0.017887 | 0.50 |

**Supplementary Table S7.**
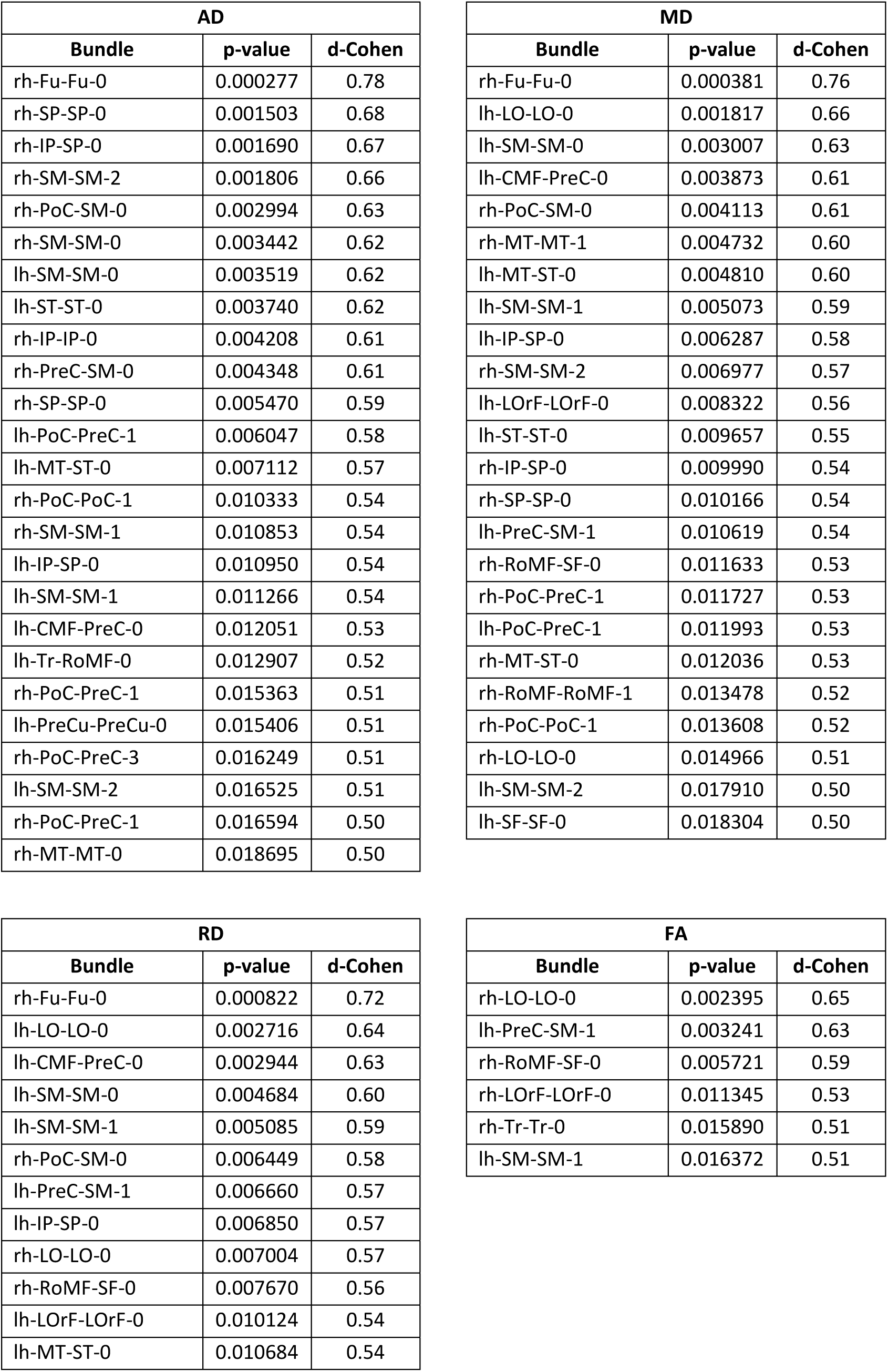
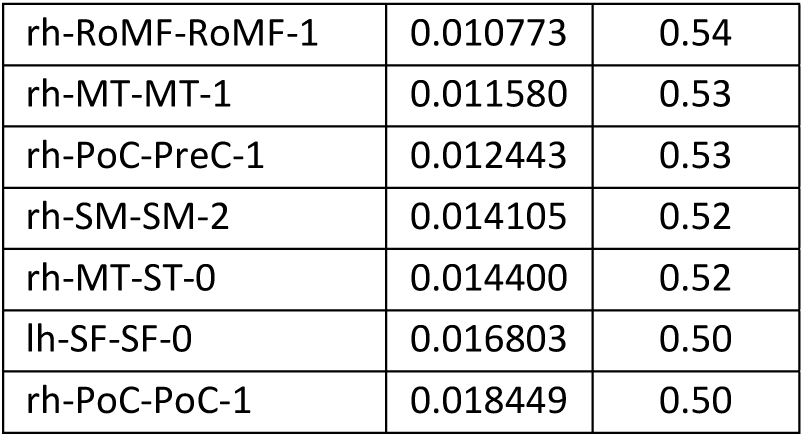
T-test statistics for the fiber bundles in SWM for diffusion tensor model metrics: AD, axial diffusivity; MD, mean diffusivity; RD, radial diffusivity; and FA, fractional anisotropy. Only bundles that demonstrate statistical significance (p-value < 0.05, uncorrected) and an effect size above 0.5 (Cohen’s d) are included. The prefixes rh and lh indicate the laterality, rh (right hemisphere) and lh (left hemisphere).

**Supplementary Table S8.** Overview of the classifiers that achieved the highest accuracy based on DWM fiber bundles for each diffusion tensor imaging (DTI) metric. For each index—AD (axial diffusivity), MD (mean diffusivity), RD (radial diffusivity), and FA (fractional anisotropy)—the most relevant fiber bundles contributing to this performance are also reported. The prefixes rh and lh indicate the laterality, rh (right hemisphere) and lh (left hemisphere).

| Report | AD | MD | RD | FA |
| --- | --- | --- | --- | --- |
| Classifier | Decision Tree | KNN | Decision Tree | KNN |
| Accuracy | 85.67 | 77.30 | 75.30 | 77.10 |
| Bundles | rh-Anterior AF<br>lh-AF<br>CC Body<br>CC Rostrum<br>lh-CG Short<br>lh-CST<br>rh-FF<br>rh-Superior MTR<br>rh-Superior PTR<br>lh-Inferior TR<br>rh-Inferior TR | rh-Anterior AF<br>rh-Posterior AF<br>rh-AF<br>rh-CG Short<br>lh-CG Long<br>lh-CST<br>lh-IFO<br>rh-IL | rh-Anterior AF<br>rh-Posterior AF<br>rh-AF<br>CC Rostrum<br>lh-CG Short<br>lh-CG Long<br>rh-IL<br>lh-Anterior TR<br>rh-Anterior TR<br>lh-Superior MTR<br>lh-Posterior TR<br>rh-Posterior TR<br>lh-Inferior TR<br>rh-UF | rh-Posterior AF<br>CC Splenium<br>lh-CST<br>rh-CST<br>rh-IFO<br>rh-IL<br>lh-Anterior TR<br>lh-Superior MTR<br>rh-Superior PTR<br>rh-UF |

**Supplementary Table S9.** Overview of the classifiers that achieved the highest accuracy based on SWM fiber bundles for each diffusion tensor imaging (DTI) metric. For each index—AD (axial diffusivity), MD (mean diffusivity), RD (radial diffusivity), and FA (fractional anisotropy)—the most relevant fiber bundles contributing to this performance are also reported. The prefixes rh and lh indicate the laterality, rh (right hemisphere) and lh (left hemisphere).

| Report | AD | MD | RD | FA |
| --- | --- | --- | --- | --- |
| Classifier | Decision Tree | Random Forest | Random Forest | XGB |
| Accuracy | 85.44 | 80.44 | 76.44 | 84.67 |
| Bundles | lh-CMF-PreC-0 | lh-CMF-PreC-0 | lh-LO-LO-0 | lh-LO-LO-1 |
|  | lh-CMF-PreC-1 | lh-Cu-Lg-0 | lh-SM-SM-0 | lh-PreC-SM-1 |
|  | lh-Cu-Lg-0 | lh-PoC-SM-0 | lh-SP-SP-0 | lh-SM-SM-1 |
|  | lh-Fu-Fu-0 | lh-SM-SM-0 | rh-LO-LO-0 | lh-Tr-SF-0 |
|  | lh-PoC-PreC-1 | rh-Fu-Fu-0 | rh-MT-MT-0 | rh-IP-LO-0 |
|  | lh-PreC-PreC-0 | rh-PoC-PreC-1 | rh-MT-MT-0 | rh-IP-SP-0 |
|  | lh-PreC-SM-0 | rh-PreC-SM-0 | rh-SF-SF-2 | rh-PoC-PreC-0 |
|  | lh-RoMF-SF-0 | rh-RoMF-RoMF-0 |  | rh-PreC-SM-0 |
|  | lh-RoMF-SF-1 |  |  | rh-RoMF-SF-0 |
|  | lh-SF-SF-0 |  |  | rh-Tr-RoMF-0 |
|  | rh-PoC-SM-0 |  |  | rh-Tr-SF-0 |
|  | rh-SP-SP-0 |  |  |  |
|  | rh-ST-ST-0 |  |  |  |
|  | rh-Tr-RoMF-0 |  |  |  |

## References

Aha DW, Bankert RL (1996) A Comparative Evaluation of Sequential Feature Selec-tion Algorithms. In: Bickel P, Diggle P, Fienberg S, et al (eds) Learning from Data, vol 112. Springer New York, New York, NY, p 199–206, 10.1007/978-1-4612-2404-419, URL http://link.springer.com/10.1007/978-1-4612-2404-419

Alhazmi FH (2020) White-matter integrity and hearing acuity decline in healthy subjects: Magnetic resonance tractography. The Neuroradiology Journal 33(3):236–243. 10.1177/1971400920913868

Ashburner J, Friston KJ (2000) Voxel-Based Morphometry—The Methods. NeuroImage 11(6):805–821. 10.1006/nimg.2000.0582

Basser PJ, Pajevic S, Pierpaoli C, et al (2000) In vivo fiber tractography using DT-MRI data. Magnetic resonance in medicine 44(4):625–632

Belkhiria C, Vergara RC, San Martín S, et al (2019) Cingulate cortex atrophy is associated with hearing loss in presbycusis with cochlear amplifier dysfunction. Frontiers in Aging Neuroscience 11:97. 10.3389/fnagi.2019.00097

Benson RR, Gattu R, Cacace AT (2014) Left hemisphere fractional anisotropy increase in noise-induced tinnitus: A diffusion tensor imaging (DTI) study of white matter tracts in the brain. Hearing Research 309:8–16. 10.1016/j.heares.2013.10.005

Bhatt JM, Lin HW, Bhattacharyya N (2016) Prevalence, severity, exposures, and treatment patterns of tinnitus in the united states. JAMA Otolaryngol Head Neck Surg 142(10):959–965. 10.1001/jamaoto.2016.1700

Bhatt JM, Bhattacharyya N, Lin HW (2017) Relationships between tinnitus and the prevalence of anxiety and depression: Tinnitus and Mood Disorders. The Laryngoscope 127(2):466–469. 10.1002/lary.26107

Breiman L (2001) Random Forests. Machine Learning 45(1):5–32. 10.1023/A:1010933404324

Catani M, Thiebautdeschotten M (2008) A diffusion tensor imaging tractography atlas for virtual in vivo dissections. Cortex 44(8):1105–1132. 10.1016/j.cortex.2008.05.004

Chen T, Guestrin C (2016) XGBoost: A Scalable Tree Boosting System. In: Proceedings of the 22nd ACM SIGKDD International Conference on Knowledge Discovery and Data Mining. ACM, pp 785–794, 10.1145/2939672.2939785

Cohen J (1988) Statistical Power Analysis for the Behavioral Sciences, 2nd edn. Routledge, New York, 10.4324/9780203771587

Cover T, Hart P (1967) Nearest neighbor pattern classification. IEEE Transactions on Information Theory 13(1):21–27. 10.1109/TIT.1967.1053964

Douglas PK, Lau E, Anderson A, et al (2013) Single trial decoding of belief decision making from EEG and fMRI data using independent components features. Frontiers in Human Neuroscience 7. 10.3389/fnhum.2013.00392

Eltabbakh A, Nada N (2023) Quantitative analysis of white matter brain changes in tinnitus patients with normal hearing: a case-controlled study with diffusion tensor imaging. Egyptian Journal of Radiology and Nuclear Medicine 54(1):101. 10.1186/s43055-023-01024-x

Ferńandez L, Veĺasquez C, Porrero JAG, et al (2020) Heschl’s gyrus fiber intersection area: a new insight on the connectivity of the auditory-language hub. Neurosurgical Focus 48(2):E7. 10.3171/2019.11.FOCUS19778

Feurer M, Hutter F (2019) Hyperparameter Optimization. In: Hutter F, Kotthoff L, Vanschoren J (eds) Automated Machine Learning: Methods, Systems, Challenges. Springer International Publishing, Cham, p 3–33, 10.1007/978-3-030-05318-51

Gonźalez Rodríguez LL, Osorio I, Cofre G. A, et al (2024) Phybers: a package for brain tractography analysis. Frontiers in Neuroscience 18:1333243. 10.3389/fnins.2024.1333243

Good CD, Johnsrude IS, Ashburner J, et al (2001) A Voxel-Based Morphometric Study of Ageing in 465 Normal Adult Human Brains. NeuroImage 14(1):21–36. 10.1006/nimg.2001.0786

Guevara M, Guevara P, Román C, et al (2020) Superficial white matter: A review on the dMRI analysis methods and applications. NeuroImage 212:116673. 10.1016/j.neuroimage.2020.116673

Guevara P, Duclap D, Poupon C, et al (2012) Automatic fiber bundle segmentation in massive tractography datasets using a multi-subject bundle atlas. NeuroImage 61(4):1083–1099. 10.1016/j.neuroimage.2012.02.071

Gunbey HP, Gunbey E, Aslan K, et al (2017) Limbic-Auditory Interactions of Tinnitus: An Evaluation Using Diffusion Tensor Imaging. Clinical Neuroradiology 27(2):221–230. 10.1007/s00062-015-0473-0

Jahromi AH, Taheri M (2017) A non-parametric mixture of Gaussian naive Bayes classifiers based on local independent features. In: 2017 Artificial Intelligence and Signal Processing Conference (AISP), pp 209–212, 10.1109/AISP.2017.8324083

Jaroszynski C, Attýe A, Job A, et al (2021) Tracking white-matter brain modifications in chronic non-bothersome acoustic trauma tinnitus. NeuroImage: Clinical 31:102696. 10.1016/j.nicl.2021.102696

Khan RA, Sutton BP, Tai Y, et al (2021) A large-scale diffusion imaging study of tinnitus and hearing loss. Scientific Reports 11(1):23395. 10.1038/s41598-021-02908-6

Knipper M, van Dijk P, Schulze H, et al (2020) The neural bases of tinnitus: Lessons from deafness and cochlear implants. Journal of Neuroscience 40(38):7190–7202. 10.1523/JNEUROSCI.1314-19.2020

Langguth B, Landgrebe M, Kleinjung T, et al (2011) Tinnitus and depression. The World Journal of Biological Psychiatry 12(7):489–500. 10.3109/15622975.2011.575178

Lebedev AV, Westman E, Van Westen GJ, et al (2014) Random Forest ensembles for detection and prediction of Alzheimer’s disease with a good between-cohort robustness. NeuroImage: Clinical 6:115–125. 10.1016/j.nicl.2014.08.023

Lee SY, Lee JY, Han, et al (2020) Neurocognition of Aged Patients With Chronic Tinnitus: Focus on Mild Cognitive Impairment. Clinical and Experimental Otorhinolaryngology 13(1):8–14. 10.21053/ceo.2018.01914

Liaw A, Wiener M (2001) Classification and regression by randomforest. Forest 23

Malcolm JG, Shenton ME, Rathi Y (2010) Filtered Multitensor Tractography. IEEE Transactions on Medical Imaging 29(9):1664–1675. 10.1109/TMI.2010.2048121

Malesci R, Brigato F, Di Cesare T, et al (2021) Tinnitus and Neuropsychological Dysfunction in the Elderly: A Systematic Review on Possible Links. Journal of Clinical Medicine 10(9):1881. 10.3390/jcm10091881

Müller AC, Guido S (2017) Introduction to machine learning with Python: a guide for data scientists, first edition. edn. O’Reilly Media, Sebastopol, CA, oCLC: 960211579

Neff P, Simões J, Psatha S, et al (2021) The impact of tinnitus distress on cognition. Scientific Reports 11(1):2243. 10.1038/s41598-021-81728-0

Ontivero-Ortega M, Lage-Castellanos A, Valente G, et al (2017) Fast Gaussian NaÃ^-^ve Bayes for searchlight classification analysis. NeuroImage 163:471–479. 10.1016/j.neuroimage.2017.09.001

Oosterloo BC, De Feijter M, Croll PH, et al (2021) Cross-sectional and longitudinal associations between tinnitus and mental health in a population-based sample of middle-aged and elderly persons. JAMA Otolaryngol Head Neck Surg 147(8):708– 716. 10.1001/jamaoto.2021.1049

Quinlan JR (1986) Induction of decision trees. Machine Learning 1(1):81–106. 10.1007/BF00116251

Raffelt DA, Tournier JD, Smith RE, et al (2017) Investigating white matter fibre density and morphology using fixel-based analysis. NeuroImage 144:58–73. 10.1016/j.neuroimage.2016.09.029

Rokach L, Maimon O (2005) Decision Trees. In: Maimon O, Rokach L (eds) Data Mining and Knowledge Discovery Handbook. Springer-Verlag, p 165–192, 10.1007/0-387-25465-X9

Román C, Guevara M, Valenzuela R, et al (2017) Clustering of Whole-Brain White Matter Short Association Bundles Using HARDI Data. Frontiers in Neuroinformatics 11:73. 10.3389/fninf.2017.00073

Rosemann S, Rauschecker JP (2023) Increased fiber density of the fornix in patients with chronic tinnitus revealed by diffusion-weighted MRI. Frontiers in Neuroscience 17:1293133. 10.3389/fnins.2023.1293133

Sarica A, Cerasa A, Quattrone A (2017) Random Forest Algorithm for the Classification of Neuroimaging Data in Alzheimer’s Disease: A Systematic Review. Frontiers in Aging Neuroscience 9:329. 10.3389/fnagi.2017.00329

Schilling K, Zhang F, Román C, et al (2025) Short association fiber tractography: key insights and surprising facts. Brain Structure and Function 230(6):97. 10.1007/s00429-025-02966-w

Schooten SV, Harel R, Ercan S, et al (2014) Applying feature selection methods on fMRI data. I: Computer Science, Medicine, URL https://www.semanticscholar.org/paper/Applying-feature-selection-methods-on-fMRI-data-Schooten-Harel/61e94ee3ffd29d41058d6b93af44c47c77165d94

Seydell-Greenwald A, Raven EP, Leaver AM, et al (2014) Diffusion Imaging of Auditory and Auditory-Limbic Connectivity in Tinnitus: Preliminary Evidence and Methodological Challenges. Neural Plasticity 2014:e145943. 10.1155/2014/145943

Shargorodsky J, Curhan GC, Farwell WR (2010) Prevalence and Characteristics of Tinnitus among US Adults. The American Journal of Medicine 123(8):711–718. 10.1016/j.amjmed.2010.02.015

Smith RE, Tournier JD, Calamante F, et al (2012) Anatomically-constrained tractography: Improved diffusion MRI streamlines tractography through effective use of anatomical information. NeuroImage 62(3):1924–1938. 10.1016/j.neuroimage.2012.06.005

Smith SM, Jenkinson M, Johansen-Berg H, et al (2006a) Tract-based spatial statistics: Voxelwise analysis of multi-subject diffusion data. NeuroImage 31(4):1487–1505. 10.1016/j.neuroimage.2006.02.024

Smith SM, Jenkinson M, Johansen-Berg H, et al (2006b) Tract-based spatial statistics: Voxelwise analysis of multi-subject diffusion data. NeuroImage 31(4):1487–1505. 10.1016/j.neuroimage.2006.02.024

Song SK, Sun SW, Ju WK, et al (2003) Diffusion tensor imaging detects and differentiates axon and myelin degeneration in mouse optic nerve after retinal ischemia. NeuroImage 20(3):1714–1722. 10.1016/j.neuroimage.2003.07.005

Stohler NA, Reinau D, Jick SS, et al (2019) A study on the epidemiology of tinnitus in the United Kingdom. Clinical Epidemiology 11:855–871. 10.2147/CLEP.S213136

Svobodová V, Profant O, Škoch A, et al (2024) The effect of aging, hearing loss, and tinnitus on white matter in the human auditory system revealed with fixel-based analysis. Frontiers in Aging Neuroscience 15:1283660. 10.3389/fnagi.2023.1283660

Torlay L, Perrone-Bertolotti M, Thomas E, et al (2017) Machine learninĝa€“XGBoost analysis of language networks to classify patients with epilepsy. Brain Informatics 4(3):159–169. 10.1007/s40708-017-0065-7

Tournier JD, Calamante F, Connelly A (2012) MRtrix: Diffusion tractography in crossing fiber regions. International Journal of Imaging Systems and Technology 22(1):53–66. 10.1002/ima.22005

Vázquez A, Ĺopez-Ĺopez N, Labra N, et al (2019) Parallel optimization of fiber bundle segmentation for massive tractography datasets. In: Proceedings of the IEEE International Symposium on Biomedical Imaging (ISBI), pp 178–181, 10.1109/ISBI.2019.8759543

Vrooman HA, Cocosco CA, Van Der Lijn F, et al (2007) Multi-spectral brain tissue segmentation using automatically trained k-Nearest-Neighbor classification. NeuroImage 37(1):71–81. 10.1016/j.neuroimage.2007.05.018

Wasserthal J, Neher PF, Hirjak D, et al (2019) Combined tract segmentation and orientation mapping for bundle-specific tractography. Medical Image Analysis 58:101559. 10.1016/j.media.2019.101559

Wehenkel M, Sutera A, Bastin C, et al (2018) Random Forests Based Group Importance Scores and Their Statistical Interpretation: Application for Alzheimer’s Disease. Frontiers in Neuroscience 12:411. 10.3389/fnins.2018.00411

Xu Q, Chai T, Yao J, et al (2025) Predominant white matter microstructural changes over gray matter in tinnitus brain. NeuroImage 312:121235. 10.1016/j.neuroimage.2025.121235

Yeh FC, Verstynen TD, Wang Y, et al (2013) Deterministic Diffusion Fiber Tracking Improved by Quantitative Anisotropy. PLOS ONE 8(11):e80713. 10.1371/journal.pone.0080713

Yi F, Yang H, Chen D, et al (2023) XGBoost-SHAP-based interpretable diagnostic framework for alzheimer’s disease. BMC Medical Informatics and Decision Making 23(1):137. 10.1186/s12911-023-02238-9

Zhang H, Schneider T, Wheeler-Kingshott CAM, et al (2012) Noddi: Practical in vivo neurite orientation dispersion and density imaging of the human brain. NeuroImage 61(4):1000–1016. 10.1016/j.neuroimage.2012.03.072

